# Evaluation of the Panbio™ rapid antigen test for SARS-CoV-2 in primary health care centers and test sites

**DOI:** 10.1101/2020.11.13.20231316

**Authors:** Oana Bulilete, Patricia Lorente, Alfonso Leiva, Eugenia Carandell, Antonio Oliver, Estrella Rojo, Pau Pericas, Joan Llobera, on behalf of COVID-19 Primary Care Research Group

**Affiliations:** Primary Health Care Research Unit, Balearic Public Health Service, Mallorca, Spain; Balearic Islands Health Research Institute (IdISBa), Mallorca, Spain; Santa Ponça Primary Health Care Center, Balearic Public Health Service, Spain; Directorate of General Health Service, Balearic Public Health Service, Spain; Microbiology Service, Son Espases University Hospital, Balearic Public Health Service, Spain

**Keywords:** COVID-19, rapid antigen test, SAR-COV-2, primary care

## Abstract

**Background:** Rapid antigen tests (Ag-RDT) are emerging as new diagnostic tools for COVID-19 and real-world evaluations are needed to establish their performance characteristics.

**Main objective:** To evaluate the accuracy of the Panbio™ Ag-RDT at primary health care (PHC) centers and test sites in symptomatic patients and close contacts, using the Reverse-Transcription Polymerase Chain Reaction (RT-PCR) test as the gold standard.

**Methods:** This was a prospective diagnostic study conducted in four PHC centers and two test sites in Mallorca, Spain. Consecutive patients older than 18 years, attending the sites for RT-PCR testing either for suggestive symptoms of infection or a close contact, were included. Two nasopharyngeal samples were collected, one for RT-PCR and the other was processed on-site using the Panbio™ rapid antigen test kit for SARS-CoV-2. The sensitivity and specificity were calculated using RT-PCR as the reference, and the predictive values using the pretest probability results for each analyzed group.

**Results:** A total of 1369 participants were included; mean age 42.5 ± 14.9 years and 54.3% women. The overall prevalence was 10.2%. Most participants (70.6%) presented within 5 days of the onset of symptoms or close contact, and more than 70% had high viral loads. The overall sensitivity was of 71.4% (95% CI: 63.1%, 78.7%), the specificity of 99.8% (95% CI: 99.4%, 99.9%), the positive predictive value of 98.0% (95% CI: 93.0%, 99.7%) and a negative predictive value of 96.8% (95% CI: 95.7%, 97.7%). The sensitivity was higher in symptomatic patients, in those arriving within 5 days since symptom onset and in those with high viral load.

**Conclusion:** Ag-RDT had relatively good performance characteristics in suspected symptomatic patients within five days since the onset of symptoms. However, our results concludes that a negative Ag-RDT in these settings must be considered as presumptive.

## Background

The COVID-19 pandemic is a significant challenge to the populations and health-care systems of countries throughout the world. The early detection of infected persons using massive and accurate testing, contact tracing, and rapid isolation are effective in slowing virus transmission. A Reverse-Transcription Polymerase Chain Reaction (RT-PCR) is currently the gold standard for diagnosis, but it has certain inconveniences, such as limited access to disposables and reagents in some regions, high cost, long processing time, and a need for specialized laboratories and trained personnel.

Rapid antigen detection tests (Ag-RDTs) were available soon after the COVID-19 pandemic began. These simple and inexpensive tests use lateral flow assays to detect proteins from the SARS-CoV-2 active infection. Nevertheless, research conducted early during the COVID-19 pandemic reported they had unsatisfactory diagnostic performance(1), especially a low sensitivity (2). A Cochrane systematic review of 8 studies of 5 different antigen tests reported an average sensitivity of 56.2% (95% confidence interval [CI]: 29.5%, 79.8%) and a mean specificity of 99.5% (95% CI: 98.1%, 99.9%)(3).

More recent studies have reported improved accuracy of Ag-RDTs, especially for patients with high viral loads, i.e. patients who present within 5 days since symptom onset(2). On 11 September, the World Health Organization (WHO) published an interim guidance that recommended use of an Ag-RDT for diagnosis of COVID-19 when the RT-PCR test is not available. The WHO recommended that an Ag-RDT test must have a sensitivity of at least 80% and a sensitivity of at least 97%, based on the gold-standard RT-PCR test (4). A patient with a positive Ag-RDT result within 5 days of symptom onset can be considered to have a SARS-CoV-2 infection because these individuals are more likely to have high viral loads. However, a negative result should be interpreted with caution, especially in patients with high pretest probability and confirmatory RT-PCR test following a negative Ag-RDT test is recommended(5).

Ag-RDTs are promising point-of-care alternatives because they can be produced at low cost in high volume, are easy to use, and provide results within 15 minutes, thus allowing quicker clinical decisions. However, the RT-PCR test remains the gold standard for diagnosis because prospective and real-world data on the performance of Ag-RDTs are currently limited (4). Nevertheless, for low- and middle-income countries that lack the resources to implement national RT-PCR testing strategies, the WHO has announced the availability of 120 million Ag-RDT kits (6). Moreover, the Bill & Melinda Gates Foundation executed separate volume guarantee agreements with Abbott and SD Biosensor for production of an Ag-RDT.

Europe is currently experiencing a second wave of the COVID-19 pandemic and there is a greater demand for diagnostic testing. Fast diagnostic testing using an inexpensive, point-of-care, easy-to-use, and rapid technique might help alleviate the burdens experienced by testing laboratories and caregivers in primary healthcare (PHC) centers and COVID-19 test sites (7). The increased demand for RT-PCR tests can cause delays in reporting of positive results and lead to delays in contact tracing, and thus have negative consequences for control of the COVID-19 pandemic.

Most previous studies that evaluated Ag-RDTs for COVID-19 examined symptomatic patients, were conducted in the setting of hospital emergency services, and examined patients who presented with moderate or severe symptoms of COVID-19. Thus, rigorous studies are needed before Ag-RDTs can be used for the diagnosis of SARS-CoV-2 in the setting of PHC centers or in the community.

The Panbio™ rapid antigen test kit for SARS-CoV-2 (Abbott Diagnostic GmbH, Jena, Germany) is a qualitative test using specimens from nasopharyngeal swabs. The manufacturer reported that the sensitivity for symptomatic patients was 93.3% overall and 98.2% in those RT-PCR cycle threshold (Ct) ≤ 33, and that the specificity was 99.4% (8).

The main aim of this study was to evaluate the performance of the Panbio™ Ag-RDT at PHC centers and test sites in symptomatic patients and close contacts, using the RT-PCR test as the gold standard.

## Methods

### Design and setting

This prospective diagnostic study was conducted in Mallorca (Balearic Islands, Spain) from October 2–25, 2020. Two testing locations (COVID-EXPRESS) that cover the city of Palma and 4 PHC centers (Santa Ponça, Alcudia, Inca, and Coll d’en Rebassa) were included.

### Study population

Individuals were invited to participate if they were older than 18 years, were not previously diagnosed with COVID-19, attended one of the above-named settings for RT-PCR testing, had symptoms suggestive of infection with referral by a general practitioner (GP), or had a close contact with another patient with an RT-PCR-confirmed infection.

All potentially eligible participants were asked to sign an informed consent document and to answer a short questionnaire that asked about the reasons for RT-PCR testing (referral by a GP due to symptoms, close contact, others); socio-demographic information (sex and age); and the presence of symptoms, type of symptoms, and number of days since symptom onset or close contact to a positive SAR-CoV-2 patient.

### SARS-CoV-2 testing

Trained nurses collected two consecutive nasopharyngeal sample swabs for the RT-PCR test and the Ag-RDT and interpreted the results of the AG-RDT on-site.

#### RT-PCR

Within 24 h of collection, one nasopharyngeal swab was sent for processing to Son Espases University Hospital, Microbiology Service without any additional information of the participants. RNA extraction was performed using the MagMAX™ Viral/Pathogen Nucleic Acid Isolation Kit (ThermoFisher) and amplification was performed using the TaqPath™ COVID-19 CE-IVD RT-PCR Kit and QuantStudio™ (ThermoFisher). The viral load was expressed as Ct for three genes (ORF, N, and S).

#### Ag-RDT

The other nasopharyngeal swab was processed on-site using the Panbio™ Ag-RDT (Abbott Diagnostic GmbH, Jena, Germany) and the results were interpreted within 15 min following the manufacturer’s instructions. This kit detects the presence of the nucleocapsid (N) protein on a membrane-based immunochromatography assay. For a positive result with the Abbot Panbio™ test, a test line must form in the result window. A visible control line is required to indicate a test result is valid. The sample from the swab is mixed with approximately 300 μl of buffer, and then 5 drops are dispensed into the device. Neither the test line nor the control line are visible in the result window prior to the specimen dispensation on the device

### Statistical analysis

To evaluate the accuracy of the Panbio™ Ag-RDT, the initial prevalence of COVID-19 was estimated as 15%, the marginal error as 5%, and the sensitivity as 90%. Thus, it was necessary to test 927 participants.

The sensitivity, specificity, and their 95% CIs were calculated using RT-PCR as the reference. Sensitivity analysis was stratified by the declared reason for performing the RT-PCR test, symptoms, days since symptom onset or exposure, and Ct-value. Predictive values and 95% CIs were estimated using the pretest probability results for each analyzed group. Means and standard deviations were used to describe population characteristics and for descriptive analysis.

All statistical calculations were performed using Stata 13 (StataCorp, College Station, TX, USA) with the Stata DIAGT module.

### Ethics and funding

This study was conducted following the Declaration of Helsinki and was approved by the Balearic Research Ethics Committee (IB 4350/20 PI on 30/09/2020) and by the Mallorca Primary Care Research Commission. Each participant was asked to sign an informed consent agreement before inclusion. This study was promoted by the Balearic Public Health Service and no external funding was received.

### Patient and public involvement

No patients were involved in setting the research question and we did not seek public engagement in the design of the laboratory aspects of the study, nor were they involved in developing plans for the design of the study. No patients were asked to advise on interpretation or writing up of results.

## Results

### Enrollment and characteristics of patients

We initially identified 1412 potentially eligible subjects visited consecutively in multiple PHC centers in Mallorca (Figure 1). Twenty-seven individuals (1.9%) declined participation, mostly because of a lack of time or anticipation of discomfort from sample collection, and we were unable to retrieve Ag-RDT results for another 16 participants. The final sample consisted of 1369 participants, their mean age was 42.5 ± 14.9 years, and 54.3% were women. The overall prevalence of COVID-19 was 10.2%, and there were 140 positive RT-PCR tests and 102 positive Ag-RDTs. We excluded 7 RT-PCR results from Panbio performance characteristics analysis (3 because of incorrect labeling that could not be recovered and 4 because of inconclusive results).

**Figure 1:**
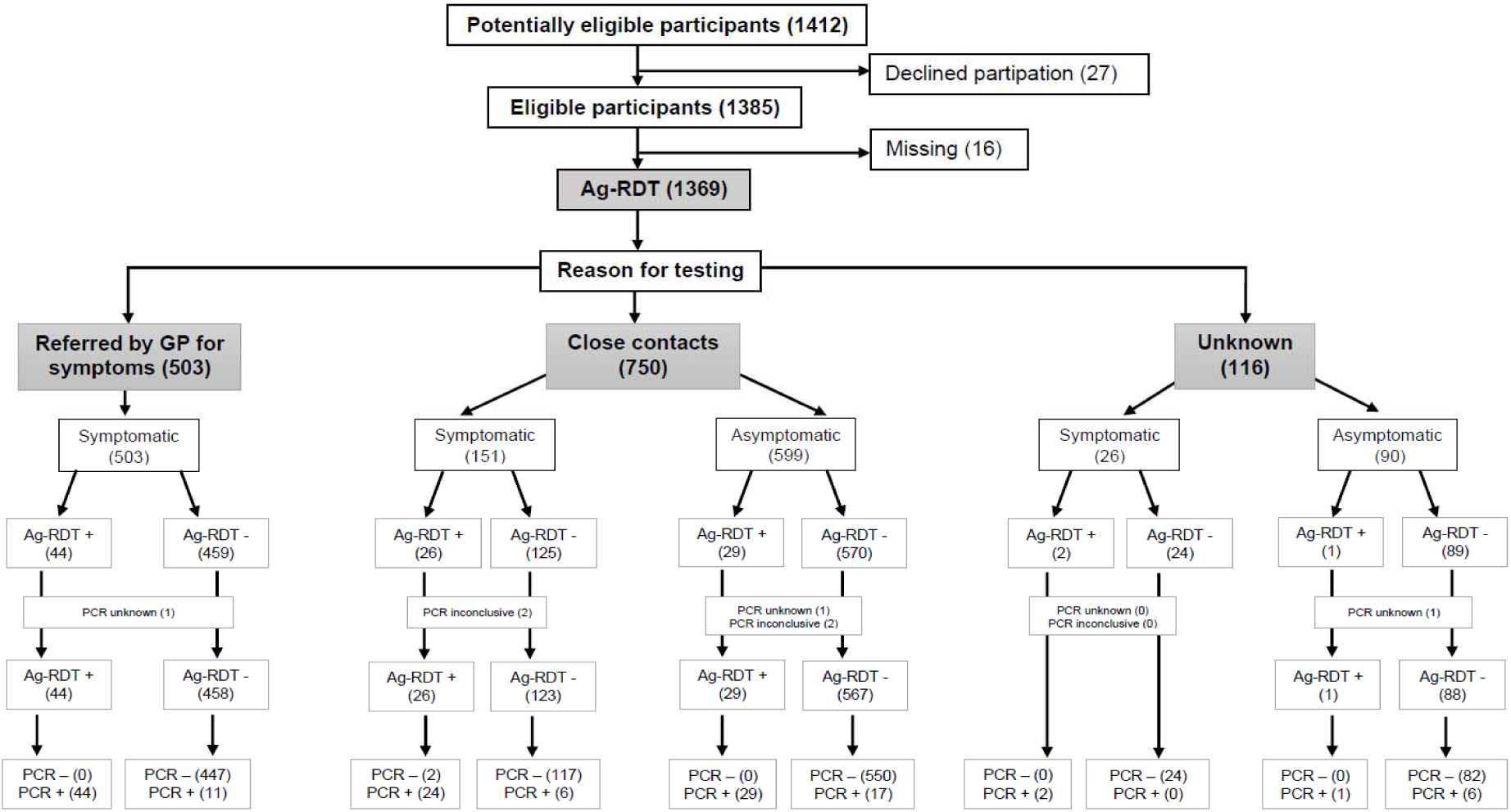
Enrollment and clinical characteristics of patients who received the RT-PCR test and the Panbio™ test.

We analyzed the characteristics of all participants (Table 1). Most appointments for RT-PCR testing were because of close contact with a confirmed positive COVID-19 individual (54.8%) or because of symptoms suggestive of COVID-19 and referral by a PHC professional (36.7%); the other 116 individuals (8.5%) were considered as unknown, because there were referred by PHC professionals without declaring the reason.

**Table 1.** Demographic and clinical characteristics of enrolled patients.

|  | **N (%)** | **RT-PCR+ (N, %)** | **Ag-RDT+ (N, %)** |
| --- | --- | --- | --- |
| **Entire sample\*** | 1369 (100%) | 140 (10.2%) | 102 (7.5%) |
| **Age (mean, SD)** | 42.5±14.9 | 41.5±14.8 | 42.8±14.0 |
| ≤20 years | 71 (5.2%) | 8 (0.6%) | 4 (0.3%) |
| 21-30 years | 272 (19.9%) | 32 (2.3%) | 19 (1.4%) |
| 31-40 years | 313 (22.9%) | 31 (2.3%) | 23 (1.7%) |
| 41-50 years | 302 (22.1%) | 31 (2.3%) | 28 (2.0%) |
| 51-60 years | 227 (16.6%) | 21 (1.5%) | 18 (1.3%) |
| 61-70 years | 132 (9.6%) | 12 (0.9%) | 6 (0.4%) |
| >70 years | 52 (3.8%) | 5 (0.4%) | 4 (0.3%) |
| **Sex** |  |  |  |
| Women | 744 (54.3%) | 71 (5.2%) | 53 (3.9%) |
| Man | 625 (45.7%) | 69 (5.0%) | 49 (3.6%) |
| **Reason for testing** |  |  |  |
| Symptoms | 503 (36.7%) | 55 (4.0%) | 44 (3.2%) |
| Close contact | 750 (54.8%) | 76 (5.6%) | 55 (4.0%) |
| Unknown | 116 (8.5%) | 9 (0.7%) | 3 (0.2%) |
| **Declaring symptoms** |  |  |  |
| Yes | 680 (49.7%) | 87 (6.4%) | 72 (5.3%) |
| No | 689 (50.3%) | 53 (3.9%) | 30 (2.2%) |
| Fever | 252 (18.4%) | 49 (3.6%) | 42 (3.1%) |
| Cough | 301 (22.0%) | 41 (3.0%) | 37 (2.7%) |
| Sore throat | 310 (22.6%) | 33 (2.4%) | 28 (2.0%) |
| Chest pain | 61 (4.5%) | 8 (0.6%) | 6 (0.4%) |
| Shortness of breath | 92 (6.7%) | 12 (0.9%) | 10 (0.7%) |
| Tiredness | 251 (18.3%) | 45 (3.3%) | 37 (3.6%) |
| Muscle/joint pain | 223 (16.3%) | 44 (3.2%) | 39 (2.8%) |
| Headache | 341 (24.9%) | 53 (3.9%) | 48 (3.5%) |
| Diarrhea | 135 (9.9%) | 9 (0.7%) | 9 (0.7%) |
| Vomiting | 50 (3.7%) | 4 (0.3%) | 4 (0.3%) |
| Loss of smell | 54 (3.9%) | 17 (1.2%) | 13 (0.9%) |
| Loss of taste | 64 (4.7%) | 19 (1.4%) | 14 (1.0%) |
| Skin involvement | 10 (0.7%) | 2 (0.1%) | 2 (0.1%) |
| Unable to move/speak | 2 (0.1%) | 1 (0.1%) | 1 (0.1%) |
| Other | 125 (9.1%) | 14 (1.0%) | 13 (0.9%) |
| Not known | 1 (0.1%) | 1 (0.1%) | 1 (0.1%) |
| **Days since SO/CC** |  |  |  |
| ≤5 | 967 (70.6%) | 101 (7.4%) | 80 (5.9%) |
| >5 | 215 (15.7%) | 19 (1.4%) | 14 (1.0%) |
| Unknown | 187 (13.6%) | 20 (1.5%) | 8 (0.5%) |
| **Viral load** |  |  |  |
| N gene(mean, SD) | 20.3±6.5 | N/A | N/A |
| S gene (mean, SD) | 21.9±6.5 | N/A | N/A |
| ORF gene (mean, SD) | 21.0±6.7 | N/A | N/A |
| N gene Ct<25 | 98 (73.1%) | 98 (73.1%) | 86 (87.8%) |
| N gene Ct=25.0-29.9 | 26 (19.4%) | 26 (19.4%) | 10 (38.5%) |
| N gene C≥30.0 | 10 (7.5%) | 10 (7.5%) | 2 (0.1%) |

RT-PCR: reverse transcription polymerase chain reaction, Ag-RDT: rapid antigen diagnostic test, SO: symptom onset, CC: close contact, Ct: RT-PCR cycle threshold.
\*4 RT-PCR results were inconclusive and 3 were unknown.

Almost half of the subjects (49.7%) reported symptoms within 7 days prior to testing, and the most frequently reported symptoms were headache (24.9%), sore throat (22.6%), cough (18.4%), and tiredness (18.3%). Most participants (70.6%) presented within 5 days of the onset of symptoms or close contact, although these data were unavailable for 13.6% of the participants. The SARS-CoV-2 N gene viral load was not obtained for 6, 7 for S gene and 8 for ORF gene, out of 140 patients with positive RT-PCR results due to sample mislabeling. The mean viral load values of the other 134 positive RT-PCR results Ct values below 25 (indicating high viral load) for all three genes (N: 20.3±6.5, S: 21.9±6.5, ORF: 21.0±6.7). We considered a Ct of 25 to 29.9 as moderate viral load and a Ct above 30 as low viral load. More than 70% of the analyzed participants (n = 98) with available Ct values had high viral loads for the N gene.

### Overall test accuracy

Our analysis of the overall performance of the Panbio™ test (Table 2) indicated a pretest prevalence of 10.2%, a sensitivity of 71.4% (95% CI: 63.1%, 78.7%), and a specificity of 99.8% (95% CI: 99.4%, 99.9%). This corresponded to a positive predictive value (PPV) of 98.0% (95% CI: 93.0%, 99.7%) and a negative predictive value (NPV) of 96.8% (95% CI: 95.7%, 97.7%).

**Table 2:** Overall sensitivity, specificity, and predictive values of the Panbio™ test.

|  | **Prevalence**<br>**(%)** | **Sensitivity**<br>**% (95% CI)** | **Specificity**<br>**% (95% CI)** | **PPV**<br>**% (95% CI)** | **NPV**<br>**% (95% CI)** |
| --- | --- | --- | --- | --- | --- |
| **Overall (N = 1362)** | 10.2% | **71.4%**<br>(63.1%, 78.7%) | **99.8%**<br>(99.4%, 99.9%) | **98.0%**<br>(93.0%, 99.7%) | **96.8%**<br>(95.7%, 97.7%) |
| **Overall: reason for testing** |  |  |  |  |  |
| GP referral for symptoms<br>(N = 502) | 10.9% | **80.0%**<br>(67.0%, 89.5%) | **100.0%**<br>(99.1%, 100%) | **100.0%**<br>(91.9%, 100%) | **97.6%**<br>(95.7%, 98.7%) |
| Close contact (N = 745) | 10.2% | **69.7%**<br>(58.1%, 79.7%) | **99.7%**<br>(98.9%, 99.9%) | **96.3%**<br>(87.4%, 99.9%) | **96.6%**<br>(95.0%, 97.8%) |
| Unknown (N = 115) | 7.8% | **33.3%**<br>(7.4%, 70.0%) | **100.0%**<br>(96.5%, 100%) | **100.0%**<br>(29.2%, 100%) | **94.6%**<br>(88.7%, 98.0%) |
| **Overall: symptoms** |  |  |  |  |  |
| Yes (N = 677) | 12.8% | **80.4%**<br>(70.5%, 88.1%) | **99.6%**<br>(98.7%, 99.9%) | **97.2%**<br>(90.3%, 99.6%) | **97.1%**<br>(95.5%, 98.3%) |
| No (N = 685) | 7.7% | **56.6%**<br>(42.3%, 70.1%) | **100.0%**<br>(99.4%, 100%) | **100.0%**<br>(88.4%, 100%) | **96.4%**<br>(94.7%, 97.7%) |

GP: general practitioner, 95%CI: 95% confidence interval, RT-PCR: Reverse-Transcription Polymerase Chain Reaction, Ag-RDT: rapid antigen diagnostic test.

Comparison of individuals referred by their GPs due to symptoms with individuals who had close contact with another patient indicated better test sensitivity (80.0%, 95% CI: 67.0%, 89.5% *vs*. 69.7%, 95% CI: 58.1%, 79.7%) and specificity (100%, 95% CI: 99.1%, 100% *vs*. 99.7%, 95% CI: 98.9%, 99.9%) in the former group. The test sensitivity was particularly poor (33.3%, 95% CI: 7.4%, 70.0%) in individuals with unknown reasons for testing. Considering all the participants reporting symptoms, the test sensitivity was similar to those with symptoms and referral by a GP (80.4%, 95% CI: 70.5%, 88.1%), but was considerably lower in asymptomatic subjects (56.6%, 95% CI: 42.3%, 70.1%). Notably, the test specificity was above 99.6% in all analyzed groups.

### Test accuracy based on days since symptom onset or close contact

As noted above, most patients (n = 963) received tests within 5 days since symptom onset or since the close contact (Table 3). Nevertheless, the overall test sensitivity for these patients was only 77.2% (95% CI: 67.6%, 84.7%), below the minimal sensitivity recommended by WHO for Ag-RDTs (80%). However, the test sensitivity was acceptable for patients who reported symptoms (n = 556; 83.1%, 95% CI: 71.9%, 90.5%) and for patients referred by their GPs for symptoms (n = 418; 86.0%, 95% CI: 71.3%, 94.2%).

**Table 3:** Accuracy of the Panbio™ test in patients tested 5 or fewer days (top) or more than 5 days (bottom) since symptom onset or close contact.

|  | **Prevalence (%)** | **Sensitivity (%; 95% CI)** | **Specificity (%; 95% CI)** | **PPV (%; 95% CI)** | **NPV (%; 95% CI)** |
| --- | --- | --- | --- | --- | --- |
| **≤5 days** |  |  |  |  |  |
| Overall (N = 963) | 10.4% | **77.2%**<br>67.6%, 84.7% | **99.7%**<br>99.0%, 99.9% | **97.5%**<br>90.4%, 99.5% | **97.4%**<br>96.0%, 98.3% |
| Overall symptomatic (N = 556) | 12.7% | **83.1%**<br>71.9%, 90.5% | **99.5%**<br>98.3%, 99.9% | **96.7%**<br>87.6%, 99.4% | **97.5%**<br>95.6%, 98.6% |
| Overall asymptomatic (N = 407) | 7.3% | **63.3%**<br>43.9%, 79.4% | **100.0%**<br>99.0%, 100% | **100.0%**<br>82.3%, 100% | **97.1%**<br>94.8%, 98, 5% |
| GP referral for symptoms (N = 418) | 10.2% | **86.0%**<br>71.3%, 94.2% | **100.0%**<br>99.0%, 100% | **100.0%**<br>90.5%, 100% | **98.4%**<br>96.4%, 99.3% |
| Overall close contacts (N = 507) | 11.0% | **71.4%**<br>57.5%, 82.3% | **99.5%**<br>98.2%, 99.9% | **95.2%**<br>82.5%, 99.1% | **96.5%**<br>94.3%, 97.9% |
| Symptomatic close contacts (N = 117) | 23.0% | **77.7%**<br>57.2%, 90.6% | **97.7%**<br>91.4%, 99.6% | **91.3%**<br>70.4%, 98.4% | **93.6%**<br>86.0%, 97.3% |
| Asymptomatic close contacts (N = 390) | 7.4% | **65.5%**<br>45.6%, 82.0% | **100%**<br>98.9%, 100% | **100%**<br>82.3%, 100% | **97.3%**<br>95.0%, 98.7% |
| **>5 days** |  |  |  |  |  |
| Overall (N = 213) | 8.9% | **73.6%**<br>48.5%, 89.8% | **100.0%**<br>98.1%, 100% | **100.0%**<br>76.8%, 100% | **97.4%**<br>93.9%, 99.0% |
| Overall symptomatic (N = 108) | 12.0% | **61.6%**<br>32.2%, 84.8% | **100.0%**<br>96.1%, 100% | **100%**<br>63.0%, 100% | **95%**<br>88.1%, 98.1% |
| Overall asymptomatic (N = 105) | 5.7% | **100.0%**<br>54.0%, 100% | **100.0%**<br>96.3%, 100% | **100.0%**<br>54.0%, 100% | **100.0%**<br>96.3%, 100% |
| GP referral for symptoms (N = 75) | 13.3% | **50.0%**<br>20.1%, 79.8% | **100.0%**<br>94.4%, 100% | **100.0%**<br>47.8%, 100% | **92.8%**<br>83.4%, 97, 3% |
| Overall close contacts (N = 135) | 6.7% | **100.0%**<br>66.3%, 100% | **100.0%**<br>97.1%, 100% | **100.0%**<br>66.3%, 100% | **100.0%**<br>97.1%, 100% |
| Symptomatic close contacts (N = 30) | 10.0% | **100.0%**<br>29.2%, 100% | **100.0%**<br>87.2%, 100% | **100.0%**<br>29.2%, 100% | **100.0%**<br>87.2%, 100% |
| Asymptomatic close contacts (N = 105) | 5.7% | **100.0%**<br>54.0%, 100% | **100.0%**<br>96.3%, 100% | **100.0%**<br>54.0%, 100% | **100.0%**<br>96.3%, 100% |

GP: general practitioner, 95%CI: 95% confidence interval, RT-PCR: Reverse-Transcription Polymerase Chain Reaction, Ag-RDT: rapid antigen diagnostic test.

The test sensitivity was also unacceptable for patients tested within 5 days since the close contact, reporting symptoms (n = 117; 77.7%, 95% CI: 57.2%, 90.6%) or not at the moment of testing (n = 390; 65.5%, 95% CI: 45.6%, 82.0%). These general trends in test sensitivity and specificity were similar when the time period was prolonged from 5 to 7 days (Supplementary Table S1). Analysis of test sensitivity according to patients’ symptoms are provided in Supplementary Table S2.

### Effect of viral load on test sensitivity

We assessed the sensitivity according to the viral gene load (Figure 2). The overall test sensitivity for patients with high viral loads of the N gene (n = 134; Ct < 25) was 87.7% (95% CI: 79.5%, 93.5%). Analysis of separate categories of patients with high viral loads indicated the test sensitivity was above 80% even in asymptomatic patients (86.2%, 95% CI: 68.3%, 96.1%). Notably, test sensitivity decreased considerably for patients with higher Ct values (lower viral loads). Analysis of S and ORF gene viral loads also indicated acceptable sensitivity of the test for patients with high viral loads, but not for patients with low viral loads (Supplementary Table S2).

**Figure 2:**
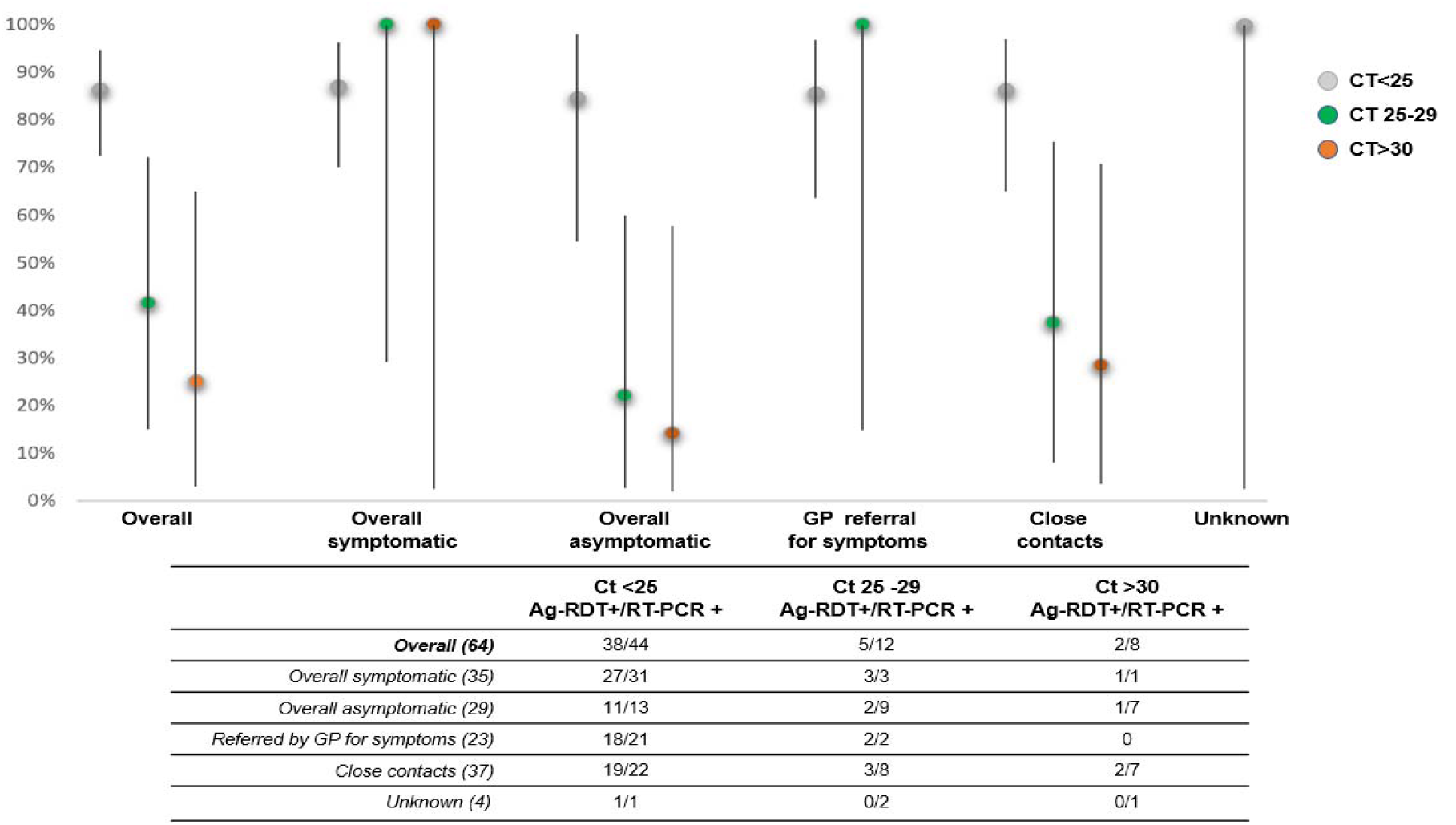
Sensitivity of the Panbio™ test in patients with different clinical status and N gene viral load.

### False-negative and false-positive results

There were false-negative Ag-RDT results in 40 of the 140 patients (28.6%) who had positive RT-PCR results. These individuals were mostly between 21 and 40 years-old (52.5%) and half of them were women. Among these 40 patients, the main reason for testing was for being in close contact with a SARS-CoV-2 confirmed patient within 5 days; 23 patients (57.3%) reported no symptoms when arriving at the testing site. Among those for whom data on N gene viral load were available (n = 36), 12 patients had high viral loads, 16 had moderate viral loads, and 8 had low viral loads.

There were false-positive Ag-RDT results in 2 of the 1222 patients (0.1%) who had negative RT-PCR results. Both of these patients were women, between 41 and 50 years-old, were tested because of close contact with a known patient, and had symptoms during the previous 5 days (headache, tiredness, and/or cough).

### Inter-observer agreement

Two independent and blinded observers, one of whom was an expert evaluator, performed visual interpretations of 68 Ag-RDT results. The interobserver agreement was 100% (Supplementary Table 4).

No side effects were reported when collecting the nasopharyngeal swabs.

## Discussion

This study was an evaluation of the Panbio Ag-RDT for SARS-CoV-2 in real-life PHC settings and test sites. Most patients visit PHC centers when they have mild or moderate symptoms of COVID-19 or after contact tracing. We found that the overall test sensitivity was 71.4% (95% CI: 63.1%, 78.7%), the sensitivity for symptomatic patients was 80.4% (95% CI: 70.5%, 88.1%), and the sensitivity for patients who reported symptoms within the previous 5 days was 83.1% (95% CI: 71.9%, 90.5%). The overall specificity was 99.8% (95% CI: 99.4%, 99.9%). Based on a prevalence of 10.2%, the PPV was 98.0% (95% CI: 93.0%, 99.7%) and the NPV was 96.8% (95% CI: 95.7%, 97.7%).

We found that test sensitivity was higher in samples that had high viral loads (Ct < 25), even in patients who were asymptomatic at the moment of testing (86.2%, 95% CI: 68.3%, 96.1%). This is an important finding because some asymptomatic individuals with SARS-CoV-2 infections (i.e., “super spreaders”) might have a substantial impact on the spreading of this virus 19 (9). The Ag-RDT provides fast results, thus facilitating early identification, rapid isolation of the patient, and early contact tracing of highly contagious cases (10,11).

### Strengths and limitations

The present study is the largest evaluation of the Panbio Ag-RDT in samples from symptomatic and asymptomatic patients in real-world context. This is a strength of our study because this is the setting where the majority of the patients with mild symptoms (up to 75%)(12) and close contacts are visited and followed-up. We incorporated COVID-19 testing into the usual care and management of patients with suspected SARS-CoV-2 infection and less than 2% of participants denied taking part in the study.

Our study has several limitations. We only assessed one type of Ag-RDT that targeted the SARS-CoV-2 N (nucleocapsid) protein, although other tests are available. Also, we only examined two types of testing scenarios —referral by a GP based on symptoms and close contact with a confirmed case — and therefore cannot make inferences about the applicability of this test in other scenarios (screening of nursing-homes, workplaces, etc.). Additional rigorous studies are needed to establish the optimal performance characteristics for Ag-RDTs that have different protein targets, are employed in different specific settings, and that have different pretest probabilities.

There is no standardized method to establishing the infectiousness of a patient with confirmed SARS-CoV-2 based on viral load. Ct values can vary among studies according to the type of test and target gene. We considered low viral load as a Ct value above 30 in patients with positive RT-PCR results (13), and assumed that such individuals can be considered non-infectious. However, our results reinforce the presence of positive relationships between a positive Ag-RDT result and high viral load in all three analyzed genes (N, S, and ORF).

### Comparation with other studies

The test sensitivity in our patients was lower than that provided by the manufacturer (93.3% overall and 98.2% in those with Ct≤ 33), but in line with previous studies. Two studies conducted in Spain that included symptomatic patients who attended a PHC center and hospital emergency wards reported the overall test sensitivity was 79.6% and the sensitivity was 86.5% in those with high viral loads (14,15). Other studies of symptomatic patients in the Netherlands and Switzerland found similar test sensitivity (85.4%, 81.0%, and 72.6%) and a positive correlation between Ag-RDT positivity and SARS-CoV-2 viral load (16,17). A study in France that examined positive and negative frozen RT-PCR samples reported a much lower overall sensitivity of only 35.3% (18); however, test sensitivity was greater in samples collected within 3 days since the onset of the symptoms and in samples from patients with high viral loads. Two preprints (non peer-reviewed studies) reported test sensitivities of 91.7% (19) and 97.1% (10). An evaluation report in Spain performed in symptomatic patients who visited hospital emergency rooms reported a sensitivity of 98.2% (20). All these previous studies reported excellent test specificity.

### Policy implications

Our results support the use of the Ag-RDT for symptomatic patients within 5 days since the onset of the symptoms in PHC setting. A positive Ag-RDT result may be considered as an active COVID-19 infection based on high specificity and PPV data. Even though the interim WHO guideline recommends use of the Ag-RDT for testing in low-income countries or when RT-PCR testing is not available, several European countries have included this new diagnostic tool as part of their national testing strategy (Belgium, France, Germany, Greece, Italy, and others) (21). Validation reports and pilot studies are being conducted to establish the diagnostic performance of these tests, and this may lead to future changes in the indications for the Ag-RDT (16,18).

Our data do not support the sole use of the Ag-RDT in asymptomatic individuals, as a negative result does not exclude the disease. We obtained an overall NPV of 96.8% in a population that had a prevalence of 10.2%, so at least 3 of 100 persons were potentially missed. Moreover, the false-negative Ag-RDT results obtained in our sample were mostly individuals who had close contact with a COVID-19 confirmed patient, were asymptomatic and most important, 12/36 participants with negative Ag-RDT result but positive RT-PCR, had high viral loads. Several other studies found that the Ag-RDT missed diagnoses in some patients with high viral loads (14,16,22). This might have a substantial impact from a public health point of view, because a negative Ag-RDT result especially in individuals with high pretest probabilities must be interpreted cautiously, and a confirmatory test should be considered(5).

There are several new approaches used to overcome the low sensitivity of the Ag-RDT, in an effort to incorporate this test as a reliable diagnostic tool for massive testing to be used for monitoring and controlling outbreaks. One approach is the complementary use of the Ag-RDT with clinical diagnostic evaluations and another approach is the use of repeated testing (23). A recent study using the enhanced epidemiological *SIDHRE-Q* model concluded that frequent Ag-RDT testing overcame the limitation of low test sensitivity, suggesting this might be an effective method to control SARS-CoV-2 transmission (24). However, future research must confirm these findings in real-world settings, as not all diagnostic tests are useful for screening(25).

Our results show that a point-of-care Ag-RDT has good performance characteristics in suspected symptomatic patients within five days since the onset of symptoms in PHC. However, a negative Ag-RDT result must be considered presumptive when the pretest probability is high, and a confirmatory test might be required. Further studies are needed to examine the accuracy of the Ag-RDT in different settings with lower pretest probabilities.

## Supporting information

STARD Checklist

## Data Availability

The anonymized database set is available at http://doi.org/10.5281/zenodo.4264502.

## Notes

### *COVID-19 Primary Care Research Group* members (alphabetical order)

Layla Aoukhiyad – *pharmacist, Servei de Salut de les Illes Balears;*

Ricardo Arcay – *medical microbiologist resident, Son Espases University Hospital Microbiology Unit;*

Javier Arranz – *general practitioner, Escola Graduada Primary Care Health Center;*

Irene Barrionuevo – *nurse, Son Espases University Hospital Emergency Unit;*

Bernardino Comas – *general practitioner, Son Espases University Hospital Emergency Unit;*

Sara Contreras – *biologist, Primary Health Care Research Unit;*

María Fernandez-Billon – *medical microbiologist resident, Son Espases University Hospital Microbiology Unit;*

J. Luis Ferrer – *nurse, Santa Ponça Primary Health Care Center;*

Pablo Fraile – *medical microbiologist resident, Son Espases University Hospital Microbiology Unit;*

Marina García – *general practitioner, Inca Primary Care Health Center;*

Carla Iglesias – *medical microbiologist resident, Son Espases University Hospital Microbiology Unit;*

Miquel Llobera – *nurse, Santa Ponça Primary Health Care Center;*

M. Consolación Méndez – *director of nursing, Mallorca Primary Health Care Directorate;*

Susana Munuera – *general practitioner, Servei de Salut de les Isles Balears;*

Victoria Pascual – *nurse, COVID Express Testing Center;*

Carlos Raduan – *general practitioner medical director, Mallorca Primary Health Care Directorate;*

Ana Requena – *nurse, Coll d’en Rabassa Primary Care Health Center;*

Antonia Roca – *general practitioner medical director, Mallorca Primary Health Care Directorate;*

Pedro Rull – *general practitioner, Son Espases University Hospital Emergency Unit;*

Silvia Vallcaneras – *nurse, Alcudia Primary Care Health Center;*

Verónica Vega – *nurse, COVID Express Testing Center.*

## Contributors

OB – protocol and data collection tools design, wrote the statistical analysis plan, validated and analyzed the data, wrote the draft and revised the paper. She is the guarantor.

PL – protocol design, implemented the whole study, monitored the whole data collection, revised the draft paper.

AL – protocol and data collection tools design, wrote the statistical analysis plan, analyzed the data, drafted and revised the paper.

EC – protocol design, implemented the study, revised the draft paper.

AO – protocol and data collection tools design, training coordinator, monitored data collection, validated the data, revised the draft paper.

ER – protocol design and data collection tools design, training coordinator, monitored data collection, validated the data, revised the draft paper.

PP – protocol and data collection tools design, validated and analyzed the data, revised the drafted paper. JL – protocol and data collection tools design, wrote the statistical analysis plan, wrote the statistical analysis plan, analyzed the data, drafted and revised the paper.

The members of the ***COVID-19 Primary Care Research Group*** design the protocol, implemented the study, monitored the data collection and revised the draft paper.

All authors approved the final version of the paper.

## Ethics

This study was conducted following the Declaration of Helsinki and was approved by the Balearic Research Ethics Committee (IB 4350/20 PI on 30/09/2020) and by the Mallorca Primary Care Research Commission. All participants had signed the informed consent before inclusion.

## Data sharing

The anonymized database set is available at http://doi.org/10.5281/zenodo.4264502.

## Transparency

The manuscript’s guarantor affirms that this manuscript is an honest, accurate, and transparent account of the study being reported; that no important aspects of the study have been omitted; and that any discrepancies from the study as planned (and, if relevant, registered) have been explained.

