## Supplemental Tables for "Evaluation of the Panbio™ rapid antigen test for SARS-CoV-2 in primary health care centers and test sites"

**Supplementary data**

**Table S1: Accuracy of the Panbio^TM^ test in patients tested 7 or fewer days (left), more than 7 days (middle), or an unknown number of days (right) since symptom onset**

|  | **Prevalence**  (%) | **≤7days** | | | | | **>7days** | | | | | **Unknown number of days** | | | | |
| --- | --- | --- | --- | --- | --- | --- | --- | --- | --- | --- | --- | --- | --- | --- | --- | --- |
|  |  | **N** | **Sensitivity**  (%, 95%CI) | **Specificity**  (%, 95%CI) | **PPV**  (%, 95%CI) | **NPV**  (%, 95%CI) | **N** | **Sensitivity**  (%, 95%CI) | **Specificity**  (%, 95%CI) | **PPV**  (%, 95%CI) | **NPV**  (%, 95%CI) | **N** | **Sensitivity**  (%, 95%CI) | **Specificity**  (%, 95%CI) | **PPV**  (%, 95%CI) | **NPV**  (%, 95%CI) |
| ***Overall*** | ≤7days =10%  >7days=9.6%  Unknown=11% | 1103 | **77%** | **99.8%** | **97.8%** | **97.4%** | 73 | **71.4%** | **100%** | **100%** | **97.1%** | 186 | **40%** | **100%** | **100%** | **93.3%** |
|  |  |  | 68.1%-84.4% | 99.3%-100% | 92.1%-99.7% | 96.3%-98.3% |  | 29%-96.3% | 94.6%-100% | 47.8%-100% | 89.8%-99.6% |  | 19.1%-63.9% | 97.8%-100% | 63.1%-100% | 88.5%-96.5% |
| ***Overall – symptomatic*** | ≤7days =13%  >7days=12%  Unknown=23% | 622 | **81%** | **99.6%** | **97%** | **97.3%** | 42 | **60%** | **100%** | **100%** | **94.9%** | 13 | **100%** | **100%** | **100%** | **100%** |
|  |  |  | 70.6%-89% | 98.7%-100% | 89.5%-99.6% | 95.6%-98.5% |  | 14.7%-94.7% | 90.5%-100% | 29.2%-100% | 82.7%-99.4% |  | 29.2%-100% | 69.2%-100% | 29.2%-100% | 69.2%-100% |
| ***Overall – asymptomatic*** | ≤7days =7.1%  >7days=6.5%  Unknown=9.8% | 481 | **67.6%** | **100%** | **100%** | **97.6%** | 31 | **100%** | **100%** | **100%** | **100%** | 173 | **29.4%** | **100%** | **100%** | **92.9%** |
|  |  |  | 49.5%-82.6% | 99.2%-100% | 85.2%-100% | 95.7%-96.8% |  | 15.8%-100% | 88.1%-100% | 15.8%-100% | 88.1%-100% |  | 10.3%-56% | 97.7%-100% | 47.8%-100% | 87.9%-96.3% |
| ***Overall – GP referral for symptoms*** | ≤7days =11%  >7days=13%  Unknown=22% | 463 | **81.6%** | **100%** | **100%** | **97.9%** | 30 | **50%** | **100%** | **100%** | **92.9%** | 9 | **100%** | **100%** | **100%** | **100%** |
|  |  |  | 68%-91.2% | 99.1%-100% | 91.2%-100% | 96%-99% |  | 6.76%-93.2% | 86.8%-100% | 15.8%-100% | 76.5%-99.1% |  | 15.8%-100% | 59%-100% | 15.8%-100% | 59%-100% |
| ***Overall – contacts*** | ≤7days =10%  >7days=7.1%  Unknown=11% | 600 | **74.2%** | **99.6%** | **95.8%** | **97.1%** | 42 | **100%** | **100%** | **100%** | **100%** | 103 | **36.4%** | **100%** | **100%** | **92.9%** |
|  |  |  | 61.5%-84.5% | 98.7%-100% | 85.7%-99.5% | 95.3%-98.3% |  | 29.2%-100% | 91%-100% | 29.2%-100% | 91%-100% |  | 10.9%-69.2% | 96.1%-100% | 39.8%-100% | 86%-97.1% |
| ***Symptomatic contacts*** | ≤7days =21%  >7days=9.1%  Unknown=50% | 136 | **79.3%** | **98.1%** | **92%** | **94.6%** | 11 | **100%** | **100%** | **100%** | **100%** | 2 | **50%** | **50%** | **50%** | **50%** |
|  |  |  | 60.3%-92% | 93.4%-99.8% | 74%-99% | 88.6%-98% |  | 2.5%-100% | 69.2%-100% | 2.5%-100% | 69.2%-100% |  | 9.1%-90.8% | 9.1%-90.8% | 9.1%-90.8% | 9.1%-90.8% |
| ***Asymptomatic contacts*** | ≤7days =7.1%  >7days=6.5%  Unknown=11% | 464 | **69.7%** | **100%** | **100%** | **97.7%** | 31 | **100%** | **100%** | **100%** | **100%** | 101 | **36.4%** | **100%** | **100%** | **92.8%** |
|  |  |  | 51.3%-84.4% | 99.1%-100% | 85.2%-100% | 95.9%-98.9% |  | 15.8%-100% | 88.1%-100% | 15.8%-100% | 88.1%-100% |  | 10.9%-69.2% | 96%-100% | 39.8%-100% | 85.7%-97% |

**or close contact with another patient.**

GP: general practitioner, 95%CI: 95% confidence interval, RT-PCR: Reverse-Transcription Polymerase Chain Reaction, Ag-RDT: rapid antigen diagnostic test.

**Table S2: Sensitivity of the Panbio^TM^ test in patients with different symptoms.**

| **Symptom** | **Sensitivity (%, 95% CI)** |
| --- | --- |
| Fever (N = 252) | 85.7% (72.7%, 94.0%) |
| Cough (N = 300) | 87.8% (73.7%, 95.9%) |
| Sore throat (N = 308) | 84.8% (68.1%, 94.8) |
| Chest pain (N = 60) | 75.0% (34.9%, 96.8%) |
| Shortness of breath (N = 91) | 83.3% (51.5%, 97.9%) |
| Tiredness (N = 251) | 80.0% (65.4%, 90.4%) |
| Muscle/joint pain (N = 223) | 86.3% (72.6%, 94.8%) |
| Headache (N = 340) | 86.7% (74.6%, 94, 5%) |
| Diarrhea (N = 135) | 88.8% (51.7%, 99.7%) |
| Vomiting (N = 50) | 100% (39.7%, 100%) |
| Loss of smell (N = 53) | 76.4% (50.1%, 93.1%) |
| Loss of taste (N = 63) | 73.6% (48.7%, 90.8%) |
| Skin involvement (N = 10) | 100% (15.8%, 100%) |
| Unable to move/speak (N = 2) | 100% (2.5%, 100%) |
| Other (N = 125) | 92.8% (66.1%, 99.8%) |
| Not known (N = 1) | 100% (2.5%, 100%) |

**Table S3: Sensitivity of the Panbio^TM^ test in patients with viral loads of the S gene (top) and ORF gene (bottom) that were high (Ct < 25), moderate (Ct = 25–29.9), or low (Ct > 30).**

|  | RT-PCR +  (N) | Ag-RDT +  (N) | **Ct <25**  Ag-RDT+/RT-PCR +  (%), 95%Ci | **Ct 25-29.9**  Ag-RDT+/RT-PCR +  (%), 95%Ci | **Ct>30**  Ag-RDT+/RT-PCR +  (%), 95%Ci |
| --- | --- | --- | --- | --- | --- |
| **S gene** | | | | | |
| **Overall** | 133 | 98 | 82/91 (90.1%)    82.0%-95.4% | 12/26 (46.2%)  26.6%-66.6% | 4/16 (25%)  7.2%-52.4% |
| *Overall symptomatic* | 84 | 69 | 59/65 (90.8%)  81%-96.5% | 8/13 (61.5%)  31.6%-86.1% | 2/6 (33.3%)  4.3%-77.7% |
| *Overall asymptomatic* | 49 | 29 | 23/26 (88.5%)  69.8%-97.6% | 4/13 (30.8%)  9.1%-61.4% | 2/10 (20%)  2.5%-55.6% |
| *Referred by GP for symptoms* | 52 | 43 | 36/39 (92.3%)  79.1%-98.4% | 6/9 (66.7%)  29.9%-92.5 | 1/4(25%)  0.6%-80.6% |
| *Close contacts overall* | 75 | 52 | 43/49 (87.8%)  75.2%-95.4% | 6/14 (42.9%)  17.7%-71.1% | 3/12 (25%)  5.5%-57.2% |
| *Symptomatic close contacts* | 30 | 24 | 21/24 (87.5%)  65.1%-97.1% | 2/4 (50%)  6.8%-93.2% | 1/2 (50%)  1.26%-98.7% |
| *Asymptomatic close contacts* | 45 | 28 | 22/25 (88%)  68.8%-97.5% | 4/10 (40%)  12.2%-73.8% | 2/10 (20%)  2.5%-55.6% |
| *Unknown overall* | 6 | 3 | 3/3 (100%)  29.2%-100% | 0/3 (0%)  0%-70.8% | 0/0 (NA) |
| **ORF gene** | | | | | |
| **Overall** | 132 | 97 | 85/95 (89.5%)  81.5%-94.8% | 10/25 (40%)  21.1%-61.3% | 2/14 (14.3%)  1.8%-42.8% |
| *Overall symptomatic* | 83 | 68 | 6168 (89.7%)  79.9%-95.8% | 6/11 (54.5%)  23.4%-83.2% | 1/4 (25%)  0.6%-80.6% |
| *Overall asymptomatic* | 51 | 29 | 24/27 (88.9%)  70.8%-94.6% | 4/14 (28.6%)  8.4%-58.1% | 1/10 (10%)  0.3%-44.5% |
| *Referred by GP for symptoms* | 52 | 43 | 38/42 (90.5%)  77.4%-97.3% | 4/7 (57.1%)  18.4%-90.1% | 1/3 (33.3%)  0.8%-90.6% |
| *Close contacts overall* | 74 | 51 | 44/50 (88%)  75.7%-95.5% | 6/15 (40%)  16.3%-67.7% | 1/9 (11.1%)  0.3%-48.2% |
| *Symptomatic close contacts* | 29 | 23 | 21/24 (87.5%)  67.6%-97.3% | 2/4 (50%)  6.8%-93.2% | 0/1(0%)  0%-97.5% |
| *Asymptomatic close contacts* | 45 | 28 | 23/26 (88.5%)  69.8%-97.6% | 4/11 (36.4%)  10.9%-69.2% | 1/8 (12.5%)  0.3%-52.7% |
| *Unknown overall* | 8 | 3 | 3/3 (100%)  29.2%-100% | 0/3 (0%)  0%-70.8% | 0/2 (0%)  0%-84.1% |

GP: general practitioner, 95%CI: 95% confidence interval, RT-PCR: Reverse-Transcription Polymerase Chain Reaction, Ag-RDT: rapid antigen diagnostic test, Ct: RT-PCR cycle threshold*.*

**Table S4: Inter-observer agreement**

|  | | **Nurse** | | **Total** |
| --- | --- | --- | --- | --- |
|  |  | **Ag-RDT +** | **Ag-RDT -** |  |
| **Expert** | **Ag-RDT +** | 5 | 0 | **5** |
|  | **Ag-RDT -** | 0 | 63 | **63** |
| **Total** | | **5** | **63** | **68** |

Ag-RDT: rapid antigen diagnostic test
